# On-site rapid molecular testing, mobile sampling teams and *eHealth* to support primary care physicians during the COVID-19 pandemic

**DOI:** 10.1101/2020.11.20.20234898

**Authors:** N.N. Cheung, S.A. Boers, S. Kiani deh Kiani, R.W. Jansen, D.O. Mook-Kanamori, L. Janssens, M.C.W. Feltkamp, A.C.M Kroes, B.C. Mourik

## Abstract

**Objectives:** We evaluated the effects of on-site rapid molecular testing at a drive-through sampling facility, deployment of mobile sampling teams and implementation of an online *eHealth* platform as supportive measures for general practitioners (GPs) during the COVID-19 pandemic.

**Methods:** An *eHealth* platform was developed that allowed GPs to either refer patients to a drive-through sampling facility or to request a home visit by a sampling team. Nasopharyngeal swab samples from patients marked as urgent (n=333) were tested immediately on-site using a GeneXpert System. Non-urgent samples (n=1,460) were sent once a day to a university hospital laboratory for routine testing. Time stamps starting from referral to the moment of test report sent were recorded to calculate the turnaround time.

**Results:** The *eHealth* platform was rapidly adopted and used by a total of 517 GPs to test 1,793 patients in a period of 13 weeks. On-site rapid molecular testing reduced the median turnaround time to 03h:41m compared to 29h:15m for routine testing. Positive SARS-CoV-2 test results were identified amongst 84/1,477 (5.7%) and 33/316 (10.4%) patients sampled at the drive-through or at home, respectively. In the age category of >80 years, 80.4% of patients were tested by a mobile sampling team.

**Conclusions:** The combination of rapid molecular testing and *eHealth* reduced the time between referral and results sent back to the GP to less than four hours. In addition, mobile sampling teams helped in reaching non-mobile, elderly patient populations with a higher prevalence of COVID-19.

## Introduction

Major global efforts have been undertaken since the emergence of novel coronavirus disease 2019 (COVID-19) to limit the spread of infection. One of the cornerstones of successful outbreak management is prompt diagnosis of patients suspected of SARS-CoV-2 infection (1). In order to achieve this, mass testing campaigns have been introduced to lower testing thresholds. Drive-through sampling facilities are often an integral part of such campaigns, because they provide an accessible, safe and efficient method for large-scale sample collection (2). An important disadvantage of such facilities however are delayed turnaround times due to transportation to nearby laboratories before the diagnostic test can be performed. In addition, drive-through facilities are often less accessible for non-mobile and/or elderly populations. In the Netherlands, these patients rely mostly on home visits by their general practitioner (GP) for sample collection. This can be time-consuming and potentially hazardous for GPs due to the already increased working hours during the pandemic combined with insufficient availability of personal protective equipment during the early phases of the pandemic (3-5). On top of these limitations, test accessibility for GPs was restricted in the Netherlanders during the early phases of the first COVID-19 wave.

To support the primary care in our region during the pandemic, we developed a system aimed at reducing diagnostic turnaround times and improved test accessibility. To this aim, we implemented an *eHealth* platform through which GPs could request a home visit by a mobile sampling team or refer patients to a drive-through sampling facility. Prior to visiting, patients could fill in their symptoms and risk profile online. During their visit at the facility, the patient’s vital functions were measured and registered in the *eHealth* system. After their visit, these data were sent back to the GP together with the test results to aid them in their further evaluation and care of the patient. The sampling facility was equipped with a mobile laboratory in which we could perform rapid molecular testing to reduce the turnaround time of patient samples marked as urgent by the GP. Results were sent directly to the GP through our *eHealth* platform to ascertain an efficient and effective route of communication.

A significant and often overlooked portion of the COVID-19 burden falls in primary care, outside of the hospital. We evaluated how implementation of these supportive measures could help GPs to better manage the increased workload of SARS-CoV-2 diagnostics during the early phase of the COVID-19 pandemic in the Netherlands, when national mass testing campaigns were still in early development.

## Materials and Methods

### Online referral of patients and eHealth consultation

The Heroku (San Francisco, United States) cloud application platform and database were used to develop an *eHealth* platform. Data was collected using questionnaires constructed with Typeform (Barcelona, Spain). Site and account access were regulated with Auth0 (Bellevue, United States). Short Message Services (SMS) communication services were used from Twilio (San Francisco, United States) and Messagebird (Amsterdam, the Netherlands).

GP’s referred patients by an online form available through an online account. Upon form completion, the GP was asked to enter the mobile telephone number of the patient, after which an SMS with a personal link to the ‘symptoms and risk profile’ questionnaire was sent to the patient. Next, a second SMS was sent to the patient, containing their personal referral code and directions to the drive-through sampling facility. After testing, a comprehensive PDF file consisting of the patient’s ‘symptoms and risk profile’ questionnaire result, vital function measurements and SARS-CoV-2 test results including interpretation from the clinical microbiology specialist were sent to back to the GP through the same *eHealth* platform.

### Location, time period and sample logistics

Our drive-through sampling and testing facility was operational between April 21^th^ and June 12^th^ 2020 at a location nearby the Hague, the Netherlands. During this period, GP’s could use our *eHealth* platform to refer patients to our facility or to request a home visit by a mobile sampling team (in collaboration with the regional Red Cross). Nasopharyngeal swab samples were transported directly to our facility after collection.

### On-site registration

A Virtual Private Network (VPN) connection with the Leiden University Medical Center (LUMC) network was available on-site. When either patients or samples arrived for registration, we entered their personal information in a specifically designed dashboard in R-software (Auckland, New Zealand). This dashboard communicated directly with GLIMS (Ghent, Belgium), the Laboratory Information System (LIS) of our Medical Microbiology department. After completion of the registration form in R, sample ID stickers from GLIMS were directly printed and placed on the test-tube prior to sampling.

### On-site sample processing

Collected swab samples were immediately processed in our mobile laboratory. Samples were vortexed and divided in a laminar flow cabinet: 1 mL was stored at −20 °C and 1 mL was either used immediately for rapid testing or temporarily stored at 4°C awaiting transportation to our clinical microbiology department.

### SARS-CoV-2 molecular testing

Urgent samples were tested on-site with the Xpress SARS-CoV-2 assay using a GeneXpert GX16-8 system (Cepheid) (6, 7). Routine samples were transported to the Medical Microbiology department of the LUMC where RNA isolation was performed using the MagNA Pure 96 (Roche, Bazel, Switzerland) and E-gen RNA amplification was done on the CFX RT-PCR platform (Bio-Rad, Hercules, United States). For routine samples, cycle-threshold (CT)-values of >35 were interpreted as negative and CT values of 30-35 were retested for confirmation using the geneLEAD VIII platform (Precision systems science, Matsudo, Japan).

### SARS-CoV-2 antigen testing

The CORIS COVID-19 antigen lateral flow assay that detects nucleoprotein antigen in respiratory samples was used (8). Briefly, 100 µL of a respiratory swab sample in GLY-medium was placed in a 15 mL tube with 4 drops of Lysis buffer and vortexed. The antigen lateral flow strip was then placed in the mixture tube for 15 minutes before read-out of the results. All result interpretations were checked by a second person.

### General Data Protection Regulation (GDPR)

This study was approved by the Institutional Review Board of the LUMC for observational studies. No patient-identifying data were stored in the cloud database. Instead, a 7-digit reference code was sent to the GP and patient by email and SMS respectively upon referral. The reference code and patient-identifying data were registered together in the GDPR-compliant LIS during on-site registration. After performing the diagnostic test, LIS sent only the reference number and the test results to the cloud application. Next, an email with a link to a PDF file containing the reference number and the test result were sent to the GP.

### ISO-15189 audit

A quality officer was assigned to ascertain that our mobile laboratory was in compliance with the International Organization for Standardization (ISO) – 15189 accreditation quality framework (9). A prospective risk analysis and an on-site internal quality audit was performed that included evaluation of all reports filed in our quality management system during the operational period.

### Statistical analysis

The Fisher’s exact test was used to determine the statistical significances in positivity rates between age categories and to determine differences in complaints between positively and negatively tested patients.

## Results

### Impact of rapid on-site molecular testing on time-to-result

A total of 1,793 patients referred by 517 GPs have been tested for SARS-CoV-2 using the *eHealth* platform (Table 1). Patients marked as urgent were tested using the on-site rapid molecular test. This concerned 199/1,477 (13.4%) of patients sampled on site and 134/316 (42.2%) of patients sampled at home.

**TABLE 1.** Number of patients and samples tested through the eHealth platform.

| Route | Urgent (n) | Pos (n) | % | Regular (n) | Pos (n) | % |
| --- | --- | --- | --- | --- | --- | --- |
| Sampled at facility (patient) | 199 | 15 | 7,5 | 1278 | 69 | 5,4 |
| Sampled at home (sample) | 134 | 11 | 8,2 | 182 | 22 | 12,1 |
| **Total** | **333** | **26** | **7,8** | **1.460** | **91** | **6,0** |

The median time from referral to result sent of patients tested either immediately on-site or by routine PCR at our clinical microbiology department is shown in Fig. 1. For urgent cases, the time period between registration and test initiation was reduced 43-fold and the time period between test initiation and report of result was reduced 6-fold compared to non-urgent cases. For patients sampled at the drive-through, this resulted in a total turnaround time of 03h:41m (95% CI: 03h:23m - 04h:06m) for urgent patients and 29h:15m (95% CI: 29h:12m - 29h:31m) for non-urgent patients. For patients sampled at home, the turnaround times were 07h:03m (95% CI: 05h:46m - 07h:36m) for urgent samples and 29h:10m (95% CI: 28h:28m – 30h:04m) for non-urgent samples.

**FIG 1.**
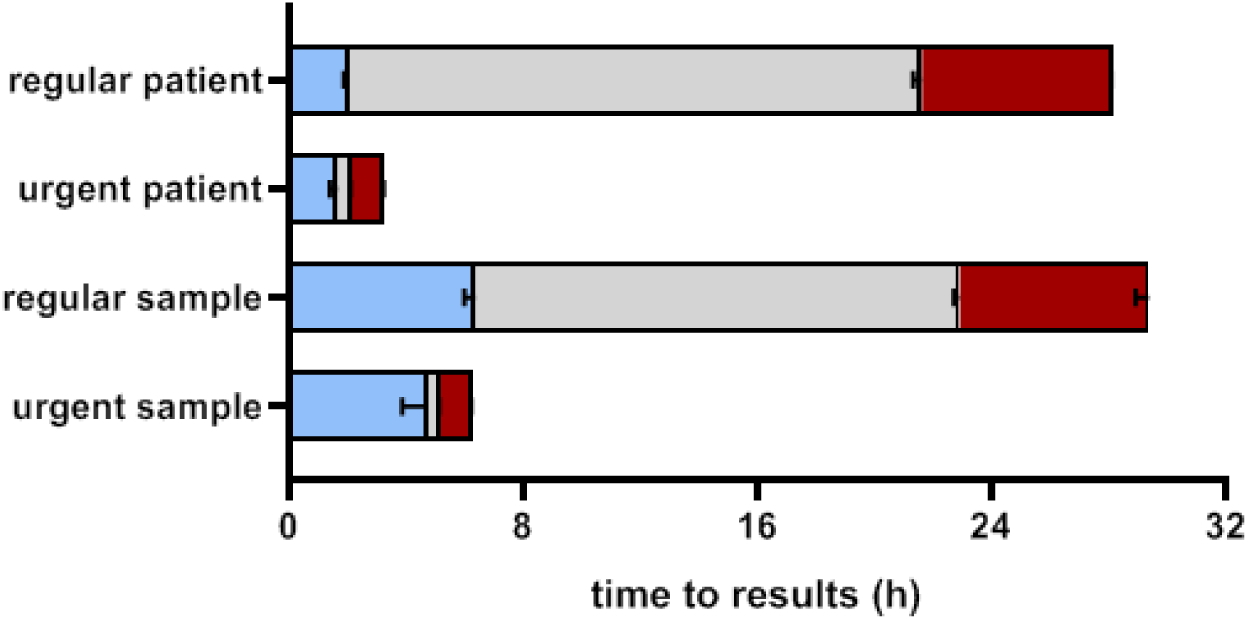
Impact of on-site rapid testing on the time-to-results. Patients that visited the sampling facility are shown as ‘patients’, while samples received from patients sampled by mobile teams are shown as ‘samples’. Blue bars show the time between referral and on-site registration, grey bars show time between on-site registration and start of diagnostics, red bars show test initiation until sending of result. Data are shown as median with 95% confidence interval. Regular patients: n=1.278, urgent patients: n=199, regular sample: n=182, urgent sample: n=134.

### Mobile sampling teams as a valuable addition to sampling facilities

Collection of nasopharyngeal swabs took place at the drive-through for 1,477 patients and at home by mobile sampling teams for 316 patients. The total number of tests performed and the percentages of positive SARS-CoV-2 test results for each age-category are shown in Fig. 2. Patients sampled at home were of a median age of 82.5 years old compared to patients sampled at the facility who had a median age of 49.7 years. In the age category of >80 years, 80.4% of patients were tested by a mobile sampling team. Patients that were sampled at home had a significantly higher percentage of positive test results of 10.4% (33/316) compared to 5.7% (84/1,477) of patients sampled at the facility (p < 0.01).

**FIG 2.**
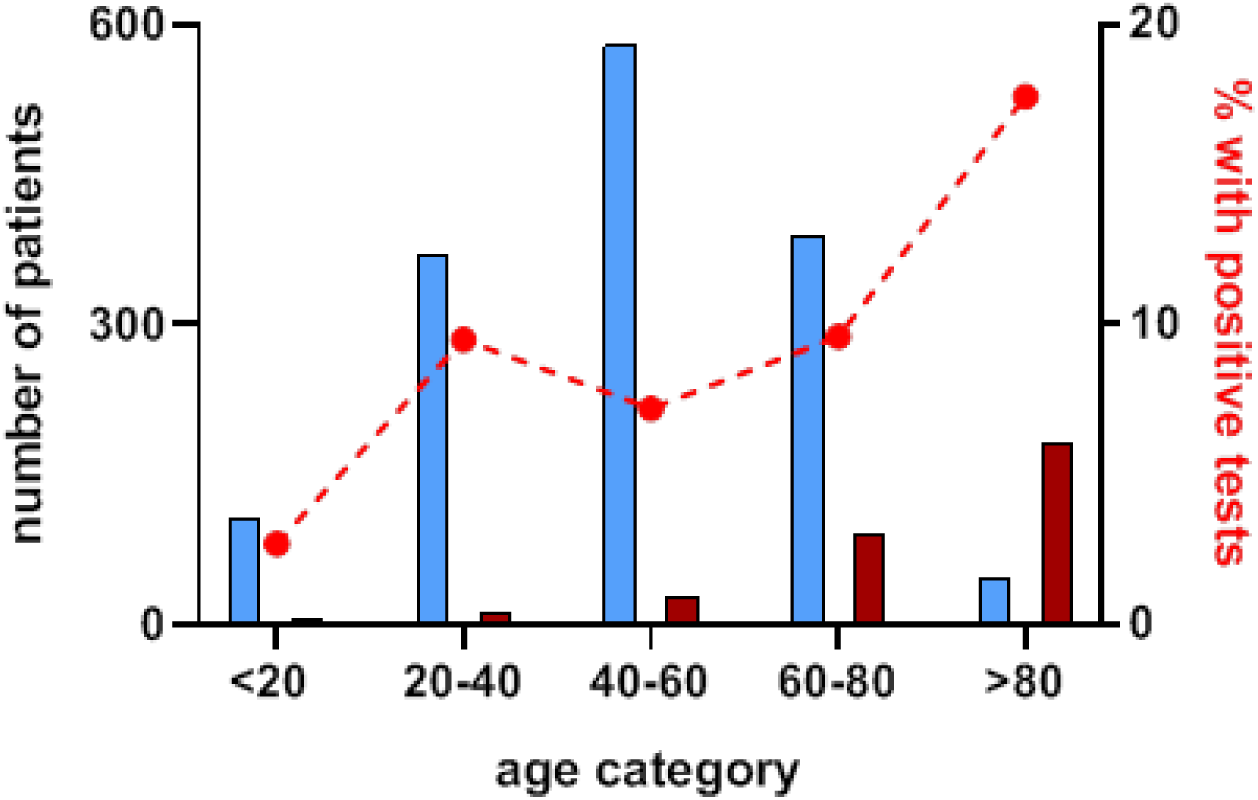
Patient age and mode of testing. Blue bars represent patients referred directly to our drive-through test facility, while red bars represent patients sampled via mobile teams. The dotted right line shows the percentage of positively tested patients per age category and is plotted against the right y-axis.

### Integrated healthcare using an eHealth platform

Combined data extracted from the ‘symptoms and risk profile’ questionnaires indicated that anosmia, fever and contact with a proven COVID-19 case within the last two weeks prior to presentation had the strongest association with a positive SARS-CoV-2 test result in our primary care population (Fig. 3).

**FIG 3.**
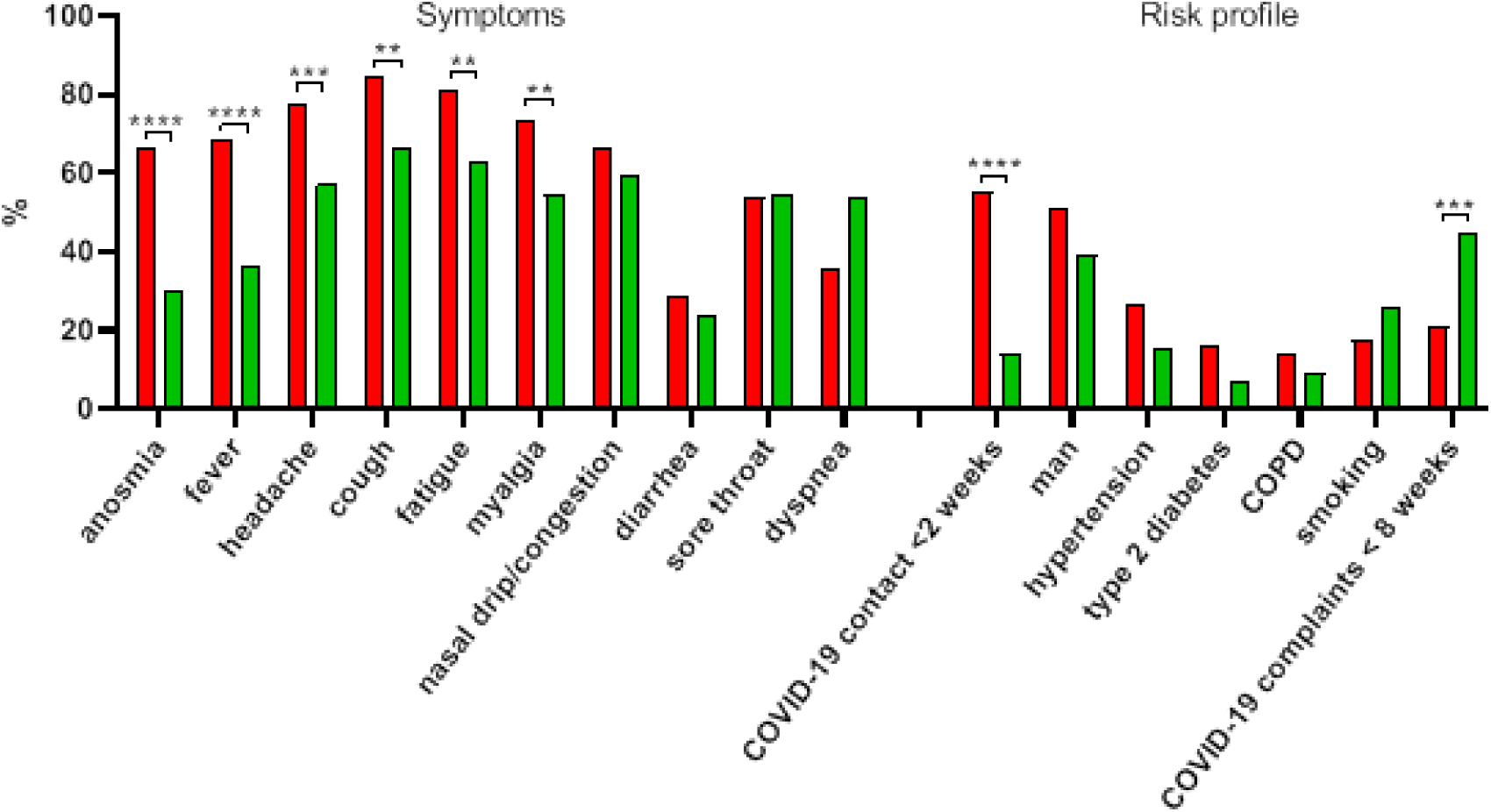
Symptoms and risk profile. Data extracted from 71 positively tested patients (red bars) and n=1.205 negatively tested patients (green bars) that completed the *eHealth* symptoms and risk profile form and provided permission to use their data. Fisher’s exact test was used to calculate significance, **** P < 0.0001, *** p < 0,001, ** p < 0,01.

### Impact of rapid antigen testing

In addition to our primary objectives, we also evaluated the use of antigen testing. This was performed on-site for n=350 samples (Fig. 4). Comparison of these results with the CT value of the routine PCR results of the exact same samples revealed that the antigen test was only able to identify positive SARS-CoV-2 samples with CT value ≤23,4. This translated to a sensitivity of 37,1% and a negative predictive value of 93,5%.

**FIG 4.**
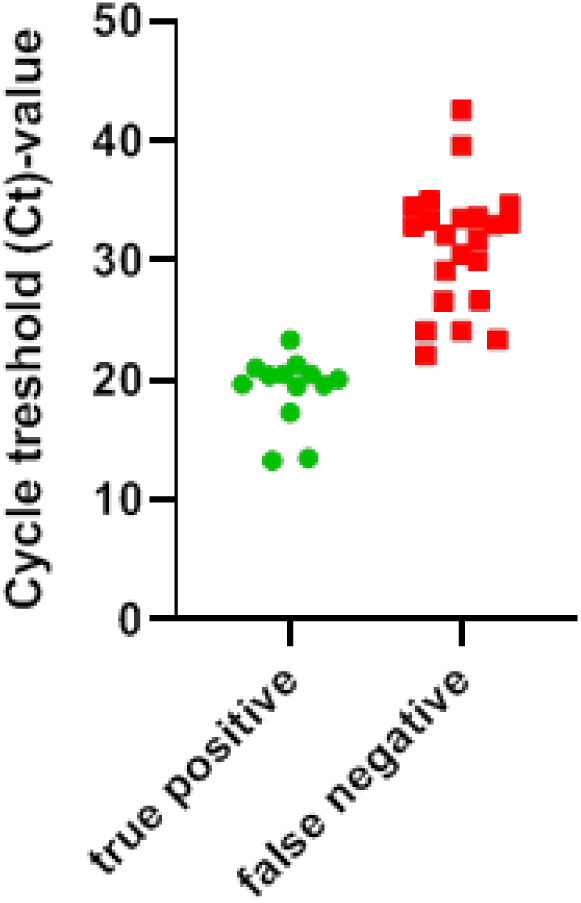
Sensitivity of rapid antigen testing. True positive antigen tests (n=13) are shown as green dots, while false negative tests (n=22) are shown as red dots. In addition, n=328 antigen tests were true negative and n=0 antigen tests were false positive (data not shown).

### Quality monitoring

An on-site ISO-15187 quality audit showed compliance on 78 points and identified 17 non-critical points that required modification. In addition 7 reports were filed in the quality management system during the operational period. None of these reports concerned direct patient care.

## Discussion

Rapid and efficient diagnostics are key-factors in coping with the current COVID-19 pandemic and should not be restricted to solely the technical qualities of a diagnostic test. Instead, efficiency should apply for the entire diagnostic process starting from sample collection to delivery of the test result to where it matters most, the patient and the treating physician. Our *eHealth* platform provided GPs with an integrated healthcare system allowing for rapid patient referrals as well as efficient collection and communication of patient’s complaints, vitals and test results. In addition, we show that on-site rapid molecular testing at a sampling facility can reduce time from referral to result sent by at least twenty-four hours compared to samples from the same facility that were tested by routine PCR in a nearby laboratory.

Mobile sampling teams complemented our facility by reaching the non-mobile and/or elderly population. These patients resided primarily in elderly homes that are known to be associated with high SARS-CoV-2 transmission-, and mortality rates (10). The importance of rapid testing in this particular population was reflected in the 3-fold higher percentage of urgent requests and a 2-fold higher percentage of positive tests compared to patients sampled at the facility. A potential selection bias where the referring GP adheres more strictly to referral guidelines before requesting a home visit cannot be excluded. Nonetheless, current mass testing campaigns in the Netherlands mainly consist of drive-throughs that are nearly inaccessible to this particular population and can therefore benefit from the addition of mobile sampling teams.

One of the advantages of performing a diagnostic test in a readily established laboratory is assurance of high quality standards and minimal risk of exposure for laboratory workers. We demonstrate that on-site diagnostics can be successfully implemented within the same quality framework as a conventional microbiology laboratory.

Next to on-site molecular testing, antigen tests are potential candidates for widespread rapid SARS-CoV-2 diagnostics. Our evaluation of one of such tests showed a limited sensitivity, which was in line with a previous evaluation (8). However, the test was simple to perform and did produce consistent results with high specificity. Also newer generations of antigen tests are entering the market with improved sensitivity. Therefore, the antigen test might prove valuable in the early diagnosis of patients with high viral loads, in low-resource settings where no molecular diagnostics are available or in surveillance settings where frequent and fast screening is desired (11, 12).

Our aim was to support the SARS-CoV-2 diagnostic process while minimizing the risks of exposure in primary care during a period of time in the Netherlands where mass testing campaigns were not yet available. Strengths of our study include the implementation of a fully integrated *eHealth* platform equipped with a data management system that allowed complete data collection and efficient, automated communication with GPs. Such platforms could be further utilized to develop algorithms for categorizing samples, e.g. in low-risk samples eligible for pooling or high-risk samples that could benefit from (on-site) rapid testing (13). Limitations might include an overly optimistic notion of the time-to-result. For example, our patient volume during the operational period consisted of a maximum of 120 patients per day, while current sampling facilities can receive up to 1500 patients each day. It is uncertain how such volumes would impact the total turnaround time.

Our data provide a first and clear picture of the impact of rapid molecular testing on time-to-result-sent outside of the hospital environment and provides a concept that could be useful for COVID-19 sampling facilities striving to reduce their turnaround time. The concept that is outlined is also relevant for the imminent widespread application of the SARS-CoV-2 antigen for rapid diagnosis of COVID-19.

## Data Availability

Data can be made available upon request.

